# Ensemble forecast of COVID-19 in Karnataka for vulnerability assessment and policy interventions

**DOI:** 10.1101/2021.08.31.21262943

**Authors:** Sashikumaar Ganesan, Deepak Subramani, Thivin Anandh, Divij Ghose, Giridhara R Babu

## Abstract

We present an ensemble forecast for Wave-3 of COVID-19 in the state of Karnataka, India, using the IISc Population Balance Model for infectious disease spread. The reported data of confirmed, recovered, and deceased cases in Karnataka from 1 July 2020 to 4 July 2021 is utilized to tune the model’s parameters, and an ensemble forecast is done from 5 July 2021 to 30 June 2022. The ensemble is built with 972 members by varying seven critical parameters that quantify the uncertainty in the spread dynamics (antibody waning, viral mutation) and interventions (pharmaceutical, non-pharmaceutical). The probability of Wave-3, the peak date distribution, and the peak caseload distribution are estimated from the ensemble forecast. Our analysis shows that the most significant causal factors are compliance to Covid-appropriate behavior, daily vaccination rate, and the immune escape new variant emergence-time. These causal factors determine when and how severe the Wave-3 of COVID-19 would be in Karnataka. We observe that when compliance to Covid-Appropriate Behavior is good (i.e., lockdown-like compliance), the emergence of new immune-escape variants beyond Sep ‘21 is unlikely to induce a new wave. A new wave is inevitable when compliance to Covid-Appropriate Behavior is only partial. Increasing the daily vaccination rates reduces the peak active caseload at Wave-3. Consequently, the hospitalization, ICU, and Oxygen requirements also decrease. Compared to Wave-2, the ensemble forecast indicates that the number of daily confirmed cases of children (0-17 years) at Wave-3’s peak could be seven times more on average. Our results provide insights to plan science-informed policy interventions and public health response.

## 1 Introduction

The COVID-19 pandemic necessitates forecasts to frame science-informed policies. An accurate forecast of the size and timing of future waves could help public health officials and governments to plan appropriate responses. An ensemble forecast by aggregating different scenarios and models makes the prediction robust and reliable.

COVID cases were reported in India from February 2020, and the first nationwide lockdown was imposed on 25 March 2020. A graded reopening started in June 2020. After that, in the state of Karnataka, the Wave-1 began gradually building up and reached a maximum daily confirmed caseload of 10K (7-day average) around 11 October 2020. Subsequently, the daily confirmed caseload reduced to less than 1K by December 2020 and stayed below 1K until mid-March 2021. From the 3rd week of March 2021, the caseload started increasing again, and a Wave-2 started raging, reaching a peak of 47.5K daily confirmed cases on 9 May 2021 [1]. Additionally, since 16 January 2021, vaccination against Covid-19 started in India with a rationing policy based on the recipient’s age [2]. Genome sequencing studies have shown that Wave-1 was caused by the B.1.1.7 (‘Alpha’) variant and Wave-2 was caused by the B.1.617.2 (‘Delta’) variant [3–7]. Despite timely NPIs (decentralized state-wise, city-wise, and community-wise lockdown), there was a widespread concern due to the high load on the medical infrastructure with reports of ICU and oxygen shortages.

Unfortunately, almost no computational model for Covid-19 spread in India predicted a Wave-2, drawing widespread flak and criticism from the Government and general public [8]. The forecast failure could have been due to the uncertainties in the spread dynamics, intervention effectiveness, virus mutation, anti-body waning, and vaccination effectiveness. As such, a comprehensive model incorporating the above uncertainties is essential to predict new waves.

Furthermore, the COVID guidelines and policies on non-pharmaceutical interventions (NPIs) and pharmaceutical interventions, including vaccination and planning, differed for Wave-1 and Wave-2 in India. Specifically, a uniform national policy was implemented during the first wave. In contrast, each state government imposed individual localized policies during the second wave. Hence, the ensemble forecast must be done at a state level to take appropriate decentralized action for containment. Here, we present an ensemble forecast of COVID-19 in Karnataka for 2021-22, focusing on predicting a new Wave-3 and quantifying its impact on vulnerable populations.

An ensemble forecast is built by aggregating predictions from different scenarios and models. Such an approach has been shown to consistently perform well in different fields, see for e.g., [9–12]. Uncertainties in human behavior, mobility, local government policies, mutation, immune response, and anti-body waning make COVID spread forecasting a challenging problem [13, 14]. An ensemble method incorporates these uncertainties as scenarios and makes the forecast more robust. Appropriate trade-offs can be made by policymakers by analyzing the probability of occurrence of each scenario being modeled.

Several authors have analysed the data of COVID Wave-2 in India and ascribed causal factors [15]. The importance of mutation in the Wave-2 and [16] the effect of immunity waning have been characterized [17]. The effect of non-pharmaceutical interventions like lockdowns [18,19] and vaccine allocation strategies [2] has been studied. Predictions for third wave based on immunity waning, emergence of an immune-escape new variant and social distancing [20] have also been made. However, no comprehensive ensemble forecast considering all known causal factors have been reported in literature. In what follows, we first describe in Sect. 2 our IISc-COVID Model, the causal control variables that we have considered and the kind of analysis that we perform. Next, we discuss our results drawing insights from the analysis done on our ensemble forecast (Sect. 3). Finally we summarize our findings and raise a call for action for effective intervention (Sec. 4).

## 2 Materials and methods

### 2.1 IISc-COVID Model

The IISc COVID model proposed in [21] is employed to compute all estimates using our in-house finite element package [22, 23]. The model consists of an unknown scalar function describing the dynamics of the infected population in a six-dimensional space. In particular, the active infected population is distributed in space, infection severity, duration of the infection, and age of the infected people. Let *T*_*∞*_ be a given final time, Ω := Ω_*x*_ ⊗ Ω_*l*_ be the spatial domain, and Ω_*l*_:= *L*_*v*_ × *L*_*d*_ × *L*_*a*_ be the internal domain. Here, *L*_*v*_, *L*_*d*_ and *L*_*a*_ denote the infection severity, duration of infection, age of the people, respectively. Then, the dynamics of the infected population *I*(*t*, **x, *l***) ∈ (0, *T*_*∞*_] × Ω_*x*_ ⊗ Ω_*l*_ is described by the population balance equation

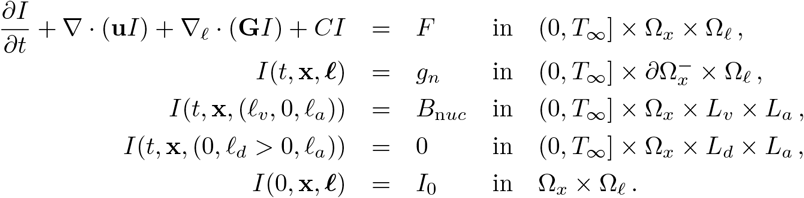

The spatial movement of the population within the spatial neighbourhood can be incorporated through **u**. The internal growth 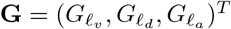 is described by

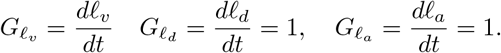

The recovery, *C*_*R*_, and infectious death, *C*_*ID*_, rates are included as *C* = *C*_*R*_ + *C*_*ID*_. The transport/mobility in the continuous model is modelled through the source term *F* defined by

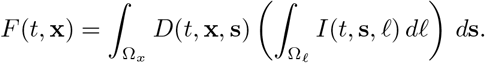

Here, the mobility function, *D*(*t*, **x, s**) : (0, *T*_*∞*_] × Ω_*x*_× Ω_*x*_ →ℝ is skew-symmetric with respect to **x** and **s** and satisfies — 1 ≤ *D*(*t*, **x, s**) ≤ 1. In particular, there will be no mobility when *D* (*t*, **x, s**) = 0 and the entire infected active population move from the place **s** to the place **x**, when *D*(*t*, **x, s**) = 1. Further, *B*_n*uc*_ and *I*_0_ are the nucleation function that quantifies the newly infected population and the initial active infected population, respectively. More details on the parameter modeling and on the numerical scheme can be found in [21]. COVID forecasting models should include antibody waning, vaccination, breakthrough infections, new immune escape variants, case-to-infection ratio (CIR) (unreported cases), social distancing, people interaction, comorbidity, travel, lockdown/unlock, etc. Eventually, all these factors affect the new infection, that is, the nucleation of COVID cases.

Let us first consider the modeling of antibody waning. Let *N* be the total number of people and *N*_*ab*_ be the number of people with antibody. Suppose there is no antibody waning, then the the susceptible ratio is defined as

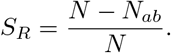

To introduce antibody waning, we use the cumulative distribution function for the Weibull distribution

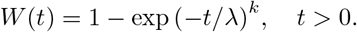

Now define the antibody waning susceptible ratio as

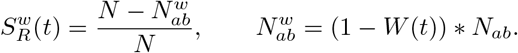

Here, *k* = 5.67 defines the shape of the curve, *λ* is the duration of antibody retention, and *t* is the time. Next, the vaccinated population must be removed from the susceptible population. Moreover, the efficacy of the vaccine should also be considered.

Suppose *N*_*v*_ is the total number of vaccinated people with 70% efficacy, then the vaccinated population is added to 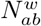, i.e.,

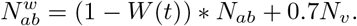

Serological surveys indicate that *CIR* varies between 20 and 60 or even more [24]. For instance, the first serological survey report of Karnataka shows *CIR* = 40 in Karnataka [25] ; that is, there are 39 unreported cases for every reported case in Karnataka during the first wave. *CIR* has to be incorporated into the model since *CIR* will significantly influence the nucleation. In our model, we included the CIR as follows:

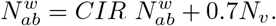

Vaccine-dependent fatality is another critical feature in our model. The straightforward approach is to define the infectious rate as a function of the vaccinated population ratio. Alternatively, vaccine-dependent fatality can also be incorporated in the nucleation when the model accounts for the severity of the infection. In particular, the newly infected but vaccinated population must be added into the model with less severity. Since one of the internal variables in our model is infection severity *l*_*v*_ and the infectious death rate is zero for *l*_*v*_ *<* 0.64, the newly infected but vaccinated population is distributed within *l*_*v*_ *<* 0.64.

Next, we discuss the modeling of new immune escape variants, which are the primary source for new waves. Suppose *N*_*ab*_(*t*) be the total antibody population on the day *t* of the introduction of a new immune escape variant. Let us assume a fraction of the antibody population, *F*_*nv*_ \**N*_*ab*_, where 0≤ *F*_*nv*_ *<* 1, is still immune to the new variant of Covid. Then, the antibody population with a new immune-escape variant is redistributed as

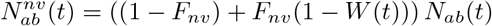

with *k* = 2.67 and *λ* = 15. Incorporating all these models, the nucleation is given by

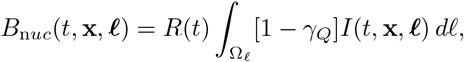

Where

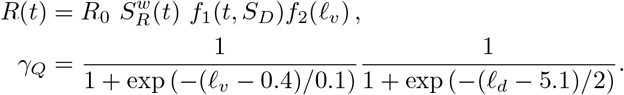

Here, *γ*_*Q*_∈ [0, 1] is the fraction of the infected population in quarantine, and it depends on testing, isolation, and comorbidity of the susceptible population. Further,

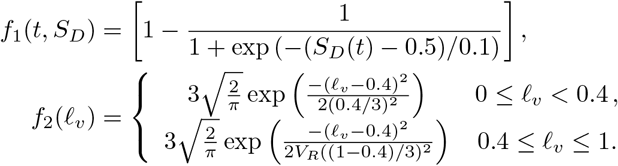

The *S*_*D*_ ∈ [0, 1] in the function *f*_1_(*t, S*_*D*_) is a social distancing parameter, where *S*_*D*_ = 1 implies a perfect social distancing and *R* → 0. Hence, *S*_*D*_ is a key parameter to forecast lockdown and unlock phases. Next, the function *f*_5_(*l*_*v*_) is used to distribute the newly infected population as a function of vaccine-dependent severity. Here, the vaccinated ratio is defined as

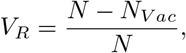

where *N*_*V ac*_ is the total number of vaccinated people.

### 2.2 Control Variables

To fit the model parameters with the dynamics of new waves and to build scenarios based on them, computations are performed from Jul 1, 2020, to Jun 30, 2022, where the actual data of the emergence of the second wave, that is, the data until Jul 4, 2021, is used. Moreover, there are many uncertainties in terms of the COVID spread [20], and a few data is not accessible to accurately fit the parameters. To account for these uncertainties, we introduce the following control variables in the model. Further, we vary these influencing parameters to build a total of 972 scenarios. Finally, an ensemble forecast is made by combining the results of these scenarios.

#### 2.2.1 Case-to-Infection Ratio (CIR)

The reported district-wise CIR values in the first serological survey for Karnataka [7, 26, 27] are used in our model. It has been reported that the average CIR in Karnataka was 40, and we denote this test case as CIR40. Further, to account for the uncertainty in the district-wise CIR estimate, we consider CIR50 as well, where the district-wise CIR is multiplied with a factor of 1.25.

#### 2.2.2 Emergence of Immune-Escape New viral Variant (IENV)

The emergence of immune-escape new viral variants is critical for the emergence of the new waves [28]. The emergence of new variants and their transmission capability depends on the circulation of the existing variants and the antibody of the infected people. Further, we can observe that the duration between the peaks across the world is between nine and thirty-five weeks. In this study, we introduce the immune-escape new viral variant after (i) eight weeks (IENV-Jul21), (ii) sixteen weeks (IENV-Sep21), and (iii) twenty-four weeks (IENV-Nov21) of the second wave (May 2021) in Karnataka. Furthermore, we consider the transmission of immune-escape new viral variants in 33% (IENV-33P), 66% (IENV-66P), and 100% (IENV-100P) of the recovered population.

#### 2.2.3 Antibody Waning (ABW)

The antibody waning could be a factor for reinfections and consequently induce new waves. A few studies show that the COVID antibody wains between five to twelve months. However, the COVID virus is new, and we do not have unambiguous evidence. Therefore, six scenarios by varying the duration of the antibody waning and the number of waning antibody populations are considered. More specifically, the antibody waning in 33% (ABW-33P), 66% (ABW-66P), and 100% (ABW-100P) of the recovered population in 150 (ABW-150) and 180 (ABW-180) days are considered.

#### 2.2.4 Vaccination Rate

Vaccination is the key to contain the spread of the virus. In the absence of treatment, preventing the spread by vaccinating the entire population is the only solution to eradicate the virus. Nevertheless, with the present vaccination rate (as of Jul 4, 2021), people in the age group of 60 and above, 45-59 Yrs., 18-44 Yrs., would be fully vaccinated by Jul 9, 21, Jul 21, 2021, and Dec 13, 2021, respectively. In addition to the availability of the vaccines, the vaccine-hesitancy is also delaying the vaccination drive. Hence to study the effect of the vaccination rate on the emergence of new waves, three scenarios, (i) present rate, which is 280K vaccinations per day (VR-280K), (ii) 50% increase in the present rate (VR-420K), and (iii) double the present rate (VR-560K), are being considered.

#### 2.2.5 COVID-appropriate behavior (CAB)

COVID-appropriate behavior (social distancing) plays a significant role in COVID spread, and it varies from region to region. Though the behavior of individuals determines the social distancing, NPIs such as lockdowns, restrictions on mass gatherings, curfew, etc., will also affect the social distancing. To study the effect of different compliance of CAB on new waves, three scenarios (i) Good compliance (Good CAB): similar to the behavior between Mar-May’21, (ii) Bad compliance (Partial CAB), and (iii) Worse compliance (No CAB), are considered.

### 2.3 Ensemble Forecast and Analysis Methods

We build an ensemble with 972 equi-probable scenarios by varying the above seven parameters. For each scenario we estimate the date of peak (if any) and the caseload at peak. A scenario without a peak is declared as a No-Wave scenario. The conditional probability of Wave-3 given a particular value for a parameter is then estimated, from only those scenarios with that chosen parameter value, as the ratio of scenarios leading to a wave 3 to the total number of scenarios. Conditional probability given two parameter values are estimated by considering only those scenarios in which those chosen parameter values are used. All statistical quantities (mean, median, confidence interval, inter-quartile range, probability distribution function and scatter) are estimated from this ensemble forecast.

## 3 Results

Our primary interest is to quantify the probability of a new COVID wave (Wave-3) in Karnataka, India, and to determine the distribution of the date and age-wise caseload of Wave-3’s peak. Most importantly, we want to determine the causal factors for a new wave and recommend suitable interventions to reduce its likelihood and impacts.

### 3.1 Causal factors for a new COVID wave in Karnataka, India

We first present the influence of the control variables (Sect. 2.2) on the emergence of a new wave and identify the significant causal factors. Among the considered 972 scenarios, Wave-3 is observed in 648 scenarios. In the event of Wave-3, the conditional probability (likelihood) of Wave-3 given a control variable is presented in Fig. 1 (a). Even though the Case-to-Infection ratio and the antibody waning could impact the emergence of new COVID waves, these factors are not very sensitive, that is, variations in these control variables are not influencing the probability of Wave-3. Nevertheless, we can observe that the variations in the new immune-escape variant, vaccination rate, and Covid-appropriate behavior significantly influence the emergence of a new COVID wave. Note that the length of the arrow indicates the sensitivity of the control variables. Moreover, we can see that the timing of the new immune-escape variant influence the probability of Wave-3 more than its reinfection percentage. Therefore, we characterize the timing of the new immune-escape variant, vaccination rate, and Covid-appropriate behavior as the primary causal factors and further analyze its influence on the Wave-3.

**Fig 1.**
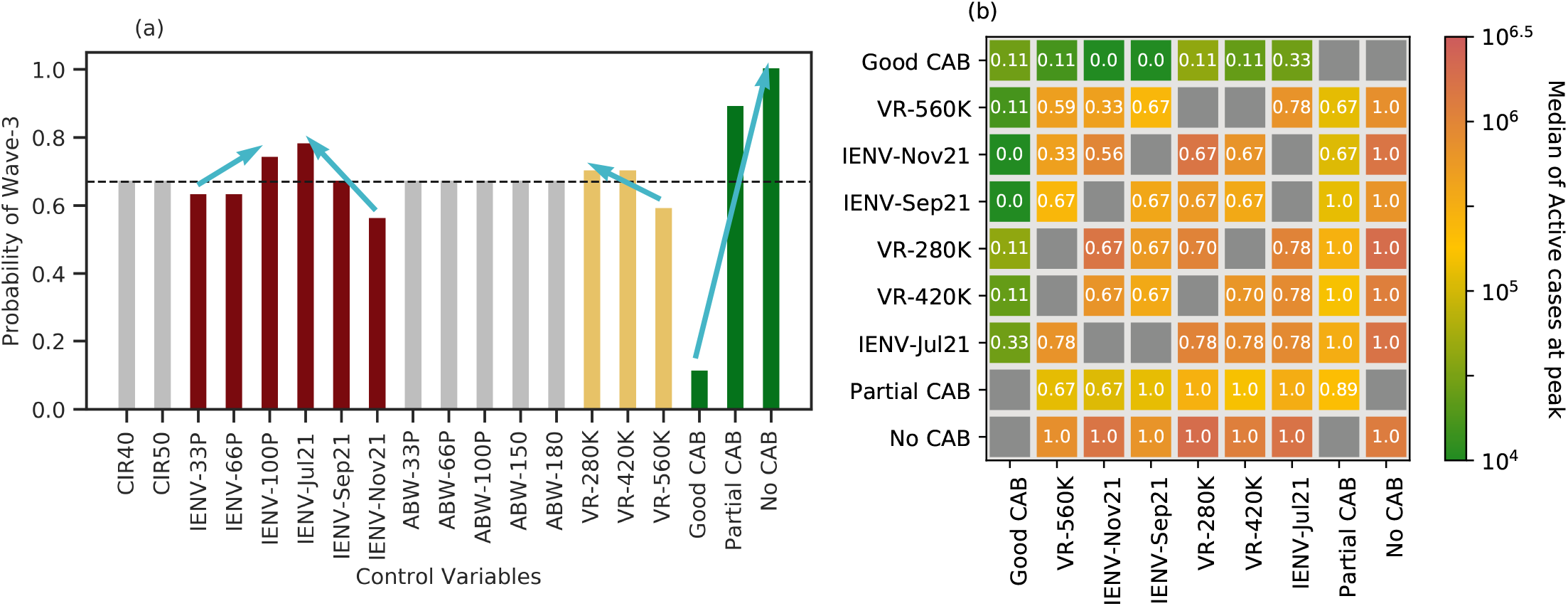
The conditional probability of Wave-3 given a control variable (a) and two control variables (b) are depicted. The values in the cells represent the conditional probability of Wave-3 given two control variables, whereas the color indicates the median of active cases at peak.

To investigate the pair-wise combined effect of these identified causal parameters on Wave-3, the conditional probability of Wave-3 given any two of these causal parameters is presented in Fig. 1 (b). The values in the cells represent the conditional probability of Wave-3 given two causal parameters, and the colors indicate the median of active cases at peak.

This analysis reveals that the CAB is the most responsible factor for the third wave. Further, no compliance of CAB guarantees the third wave even with a doubled vaccination rate and more active cases at the peak of Wave-3. On the contrary, good compliance of CAB and doubling the present vaccination rate effectively reduce the probability of Wave-3 and the number of active cases in the event of Wave-3.

### 3.2 Wave-3 peak date and active caseload

The quantities of interest for policy-making are the peak date and active caseload at peak. Now, we proceed to analyze these two quantities in different scenarios corresponding to the primary causal factors identified above. Fig. 2 shows a scatter plot between the date of Wave-3’s peak and active caseload at peak for different scenarios. A special axis entry NW is used to depict the scenarios that do not show a new wave. The arrangement and color of scatter markers in the figure help us visualize the joint-effect of adherence to Covid-appropriate behavior by the public, IENV emergence and re-infection fraction, vaccination rate, and CIR in the population on caseload and date of peak. Each panel (a-i) in Fig. 2 shows only those scenarios in which the variable mentioned in the column (CAB) and the row (IENV emergence time) are active. For example, panel (a) shows all the scenarios where the CAB is set to “Good” level and IENV emergence time is set to July 2021. Within the panel, each point represents a particular scenario. Each scenario is colored by the vaccination rate, marked by a marker that represents the IENV re-infection fraction, and the border is colored by the CIR.

**Fig 2.**
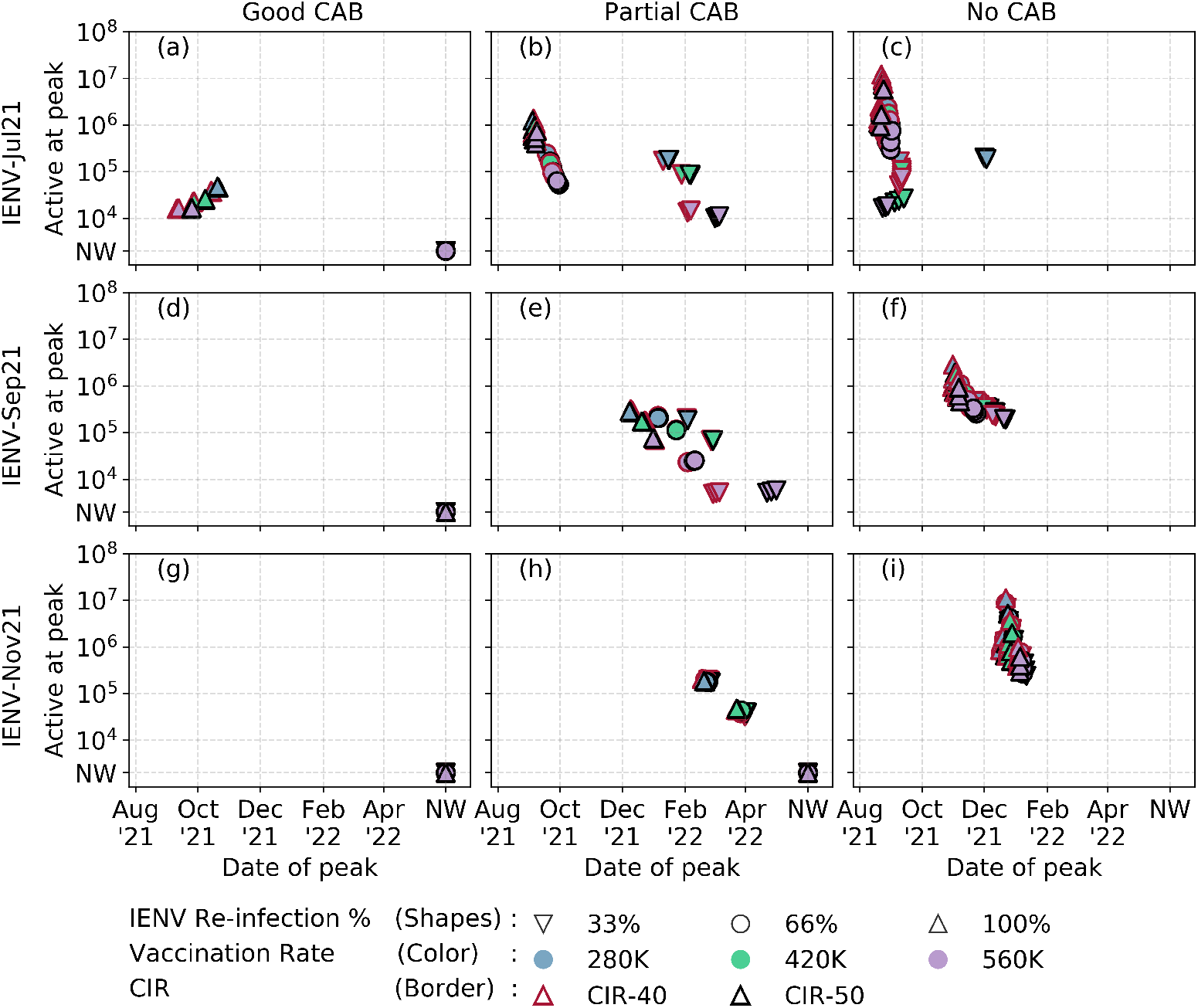
Scatter plot of active cases at the peak of Wave-3 v/s the date of peak for different scenarios of CAB and timing of the immune-escape variant with different vaccine rates.

First, we see that when Good CAB is active (panels a,d,g) as opposed to the other two values for CAB, the active case at the peak is lower. Significantly, scenarios with Good CAB and IENV emergence time beyond September 2021 (panels d,g) do not see a new wave. In panel (a), we see a peak only when 100% IENV re-infection occurs, and the other two re-infection fractions do not lead to a new wave. This result shows that Good CAB is the primary intervention strategy for preventing COVID waves, a well-known fact. However, in reality, the financial and livelihood requirements to re-open economies will result in a relaxation of CAB, and only a Partial CAB is likely to be followed. In those scenarios with partial CAB (panels b,e,h), if an immune escape new variant emerges in July 2021 or September 2021, Wave-3 is inevitable in Karnataka. Moreover, we see that vaccination rate is the key governing factor that reduces the active caseload at peak. Crucially, reaching a vaccination level of 580k per day in Karnataka will ensure no Wave-3 if no immune escape new variant starts spreading in the population before November 2021. A higher CIR (meaning lower reported cases during Wave-2) will result in a delayed peak but at the same active caseload as seen by the clustering among the same color and marker shapes.

To complete the analysis, we look at the scenarios with no CAB. Under these scenarios, we expect the highest levels of the caseload at peak and an assured emergence of new COVID waves in Karnataka. Fortunately, we see that vaccination reduces the caseload but doesn’t prevent the inevitable Wave-3. The re-infection fraction significantly affects the caseload in the No CAB, IENV-Jul’21 scenarios (panel c). We see wide distribution in the caseload at the peak but a narrow distribution in the peak date.

To further understand Wave-3 properties from the perspective of vaccination, Fig. 3 shows the probability distribution function of peak date and active cases at peak for different vaccination rates. Here, we only look at the scenarios resulting in a Wave-3 and ignore those that do not lead to a Wave-3. We see that almost all the distribution curves are multi-modal, indicating the cross effects of CAB and IENV re-infection rates. It is evident from Fig. 3 that the emergence of a new immune-escape variant is likely to be followed by a third wave within 45 to 60 days. In most cases, a higher vaccination rate helps to bring down the active caseload at peak. Doubling the daily vaccination rate (VR-560K, Fig. 3(c),(f)) results in fewer scenarios with Wave-3 despite the emergence of an immune-escape new variant in November.

**Fig 3.**
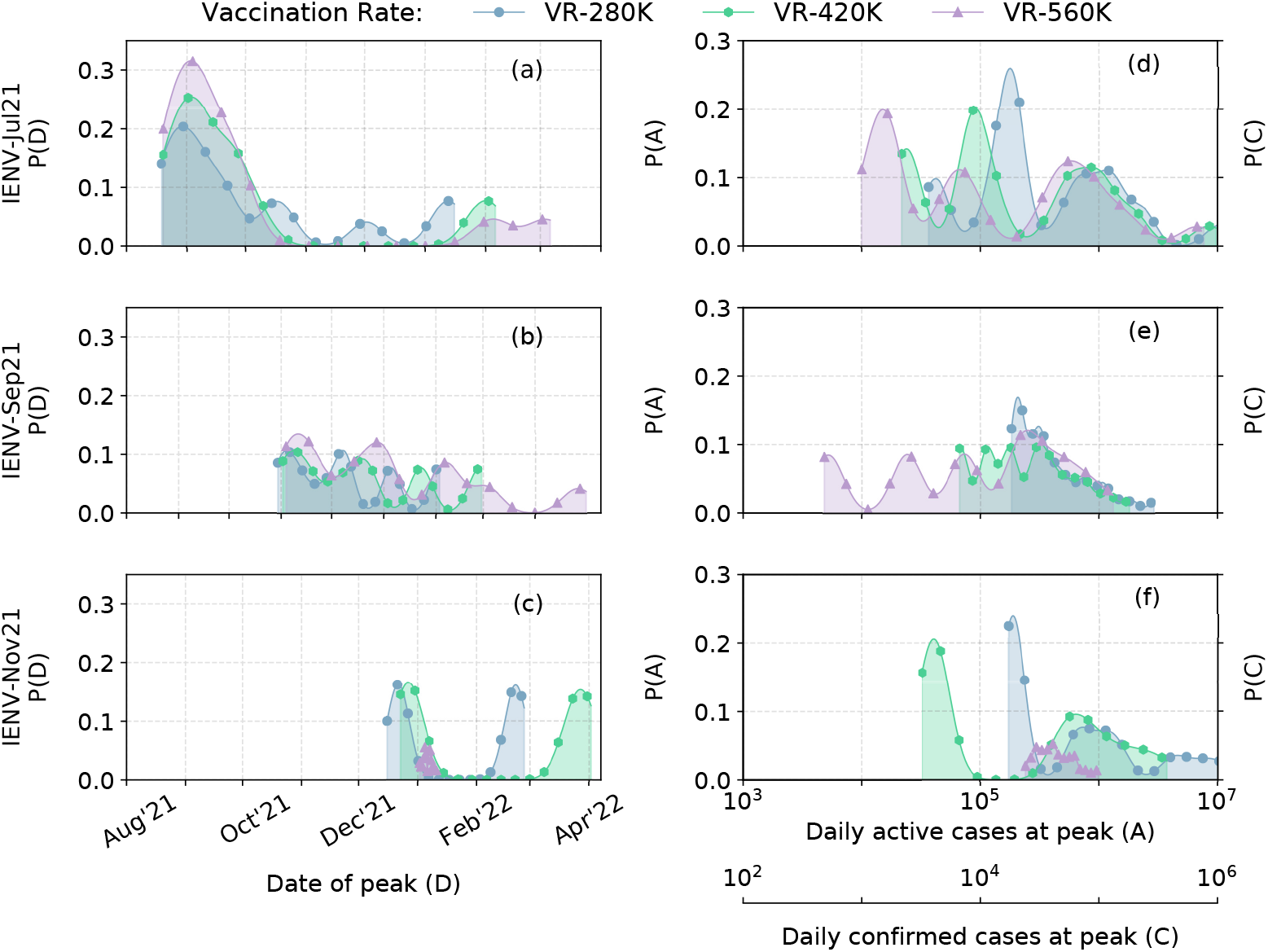
The probability of the predicted date of peak, the number of active cases and the number of confirmed cases on the day of peak for different vaccination rates and timing of emergence of immune-escape new variants, i.e., in Jul’21, Sep’21 and Nov’21.

Table 2 shows a summary of the median peak date, inter-quartile range of peak date and the minimum and maximum active caseload during this period of inter-quartile range for scenarios with specific combinations of IENV emergence time and vaccination rates. This table can be used as a ready reference for policy making.

**Table 1.** Statistics for total active cases and the date of peak at various vaccine rates, and COVID-appropriate behavior. The first value in the cell is the the median date of peak and the mean value of active cases on the median date of peak, followed by the first and third quartile values for the date of peak, and the minimum and maximum number of active cases in the inter-quartile range.

| Vaccination rate<br>per day | Emergence of<br>IENV | Good CAB | Partial CAB | No CAB |
| --- | --- | --- | --- | --- |
| 280K | Jul'21 | 15-Oct'21 (36K)<br>14-Oct'21, 21-Oct'21<br>36K – 48K | 21-Sep'21 (173K)<br>05-Sep'21, 10-Jan'22<br>144K – 207K | 27-Aug'21 (451K)<br>22-Aug'21, 10-Sep'21<br>451K – 2M |
|  | Sep'21 | No Wave-3 | 05-Jan'22 (220K)<br>10-Dec'21, 02-Feb'22<br>207K – 229K | 08-Nov'21 (674K)<br>02-Nov'21, 24-Nov'21<br>527K – 766K |
|  | Nov'21 | No Wave-3 | 21-Feb'22 (189K)<br>19-Feb'22, 24-Feb'22<br>179K – 197K | 22-Dec'21 (7M)<br>20-Dec'21, 24-Dec'21<br>616K – 11M |
| 420K | Jul'21 | 27-Sep'21 (23K)<br>26-Sep'21, 08-Oct'21<br>23K – 26K | 23-Sep'21 (119K)<br>06-Sep'21, 28-Jan'22<br>89K – 134K | 29-Aug'21 (1M)<br>23-Aug'21, 08-Sep'21<br>370K – 1M |
|  | Sep'21 | No Wave-3 | 23-Jan'22 (114K)<br>23-Dec'21, 26-Feb'22<br>112K – 117K | 13-Nov'21 (479K)<br>05-Nov'21, 02-Dec'21<br>377K – 529K |
|  | Nov'21 | No Wave-3 | 27-Mar'22 (38K)<br>24-Mar'22, 31-Mar'22<br>37K – 43K | 27-Dec'21 (1M)<br>26-Dec'21, 29-Dec'21<br>460K – 3M |
| 560K | Jul'21 | 12-Sep'21 (16K)<br>11-Sep'21, 24-Sep'21<br>15K – 16K | 25-Sep'21 (79K)<br>08-Sep'21, 04-Feb'22<br>57K – 88K | 28-Aug'21 (225K)<br>23-Aug'21, 31-Aug'21<br>17K – 656K |
|  | Sep'21 | No Wave-3 | 03-Feb'22 (23K)<br>02-Jan'22, 03-Mar'22<br>23K – 25K | 17-Nov'21 (340K)<br>06-Nov'21, 11-Dec'21<br>288K – 406K |
|  | Nov'21 | No Wave-3 | No Wave-3 | 05-Jan'22 (529K)<br>04-Jan'22, 08-Jan'22<br>296K – 792K |

**Table 2.**
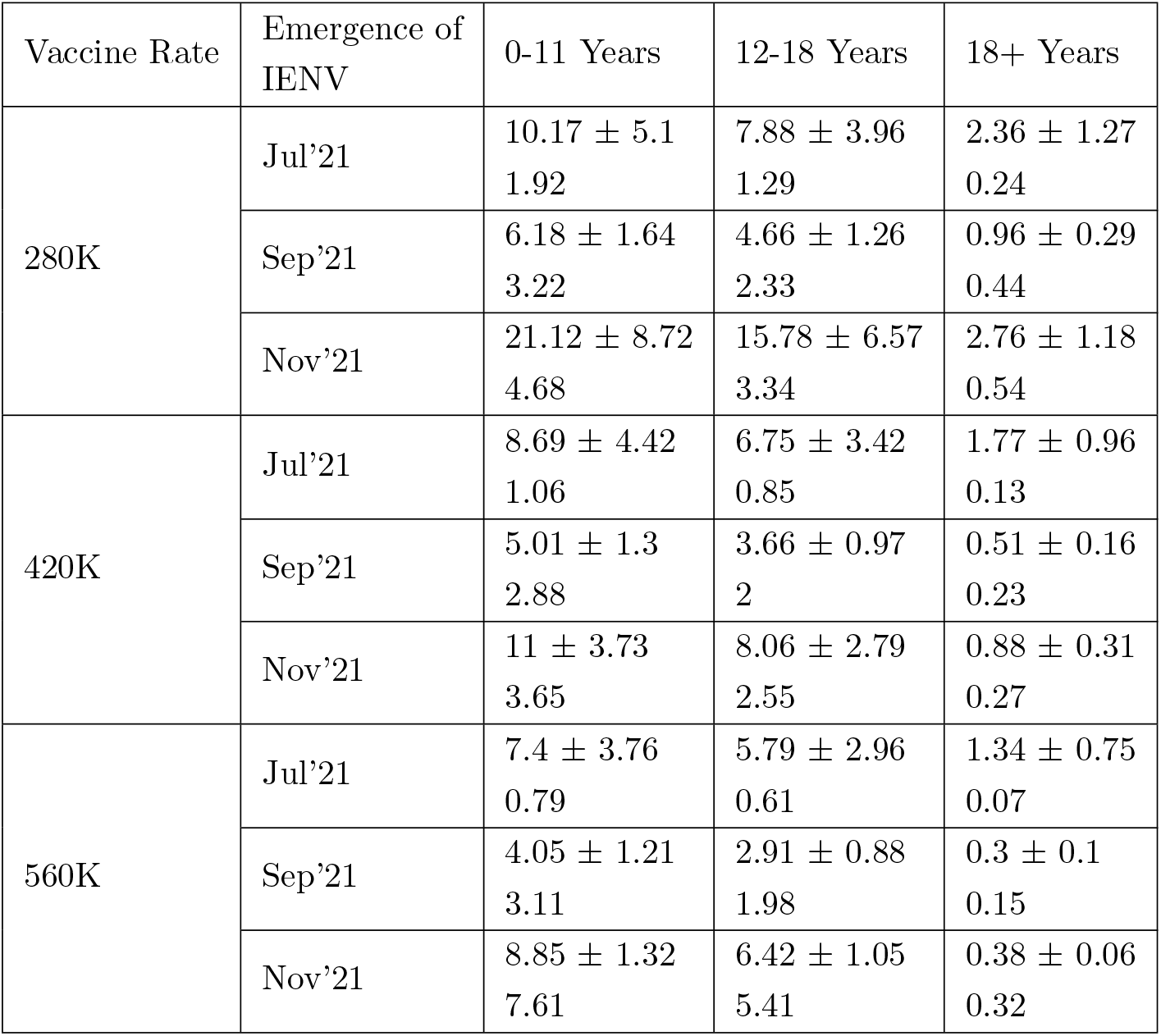
Predicted impact of Wave-3 on different age groups with different vaccination rate. The first value in the cell is the mean with the confidence interval of ratios of the 7-day average of confirmed cases at Wave-3 peak and Wave-2 peak, followed by the median.

| Vaccine Rate | Emergence of IENV | 0-11 Years | 12-18 Years | 18+ Years |
| --- | --- | --- | --- | --- |
| 280K | Jul'21 | $10.17 \pm 5.1$<br>1.92 | $7.88 \pm 3.96$<br>1.29 | $2.36 \pm 1.27$<br>0.24 |
| | Sep'21 | $6.18 \pm 1.64$<br>3.22 | $4.66 \pm 1.26$<br>2.33 | $0.96 \pm 0.29$<br>0.44 |
| | Nov'21 | $21.12 \pm 8.72$<br>4.68 | $15.78 \pm 6.57$<br>3.34 | $2.76 \pm 1.18$<br>0.54 |
| 420K | Jul'21 | $8.69 \pm 4.42$<br>1.06 | $6.75 \pm 3.42$<br>0.85 | $1.77 \pm 0.96$<br>0.13 |
| | Sep'21 | $5.01 \pm 1.3$<br>2.88 | $3.66 \pm 0.97$<br>2 | $0.51 \pm 0.16$<br>0.23 |
| | Nov'21 | $11 \pm 3.73$<br>3.65 | $8.06 \pm 2.79$<br>2.55 | $0.88 \pm 0.31$<br>0.27 |
| 560K | Jul'21 | $7.4 \pm 3.76$<br>0.79 | $5.79 \pm 2.96$<br>0.61 | $1.34 \pm 0.75$<br>0.07 |
| | Sep'21 | $4.05 \pm 1.21$<br>3.11 | $2.91 \pm 0.88$<br>1.98 | $0.3 \pm 0.1$<br>0.15 |
| | Nov'21 | $8.85 \pm 1.32$<br>7.61 | $6.42 \pm 1.05$<br>5.41 | $0.38 \pm 0.06$<br>0.32 |

### 3.3 Effect of Wave-3 on different age-groups

The data until 10 June 2021 shows that 4.2%, 5%, 53.2%, 23% and 14.645 % of the people in the age groups 0–11, 12–17, 18–44, 45–59, and above 60 years were infected with COVID-19. However, India started an age-dependent vaccination drive for 18+ population only. As such, for Wave-3, we expect a higher contribution to the daily confirmed caseload from children (0–17 years). To estimate this impact on different age groups, we analyze the age-wise daily confirmed caseload at Wave-3 peak and compare it with the corresponding numbers at Wave-2. Table. 2 shows the statistics of the ratio between the 7-day average of daily confirmed cases of Wave-3 and Wave-2 peaks. The first line in each cell shows the ensemble mean with the 95% confidence interval and the second line shows the ensemble median. We see that the children (0-17 years) are the worst affected due to unavailability of vaccine. In the worst case,with VR-280K and IENV-Nov21, the daily confirmed cases of children (0–11 years) at the Wave-3 peak will be on an average twenty times more than that of the daily confirmed cases at Wave-3 peak. On an average, the impact of Wave-3 on children (0–17 years) would be seven times more than that of Wave-2.

### 3.4 Active Cases During May’21 - Jun’22

Fig. 4 shows the ensemble forecast of active cases in Karnataka from May 2021 to June 2022. Each panel shows the scenarios given a pair of CAB and IENV emergence-time control variables. The colored solid line is the mean value of the ensemble of active cases, and the shaded zone is the region of uncertainty (each color corresponds to a particular vaccination rate). The rows in Fig. 4 represent the good, partial, and no CAB, whereas the columns represent the emergence of IENV in Jul’21, Sep’21, and Nov’21, respectively.

**Fig 4.**
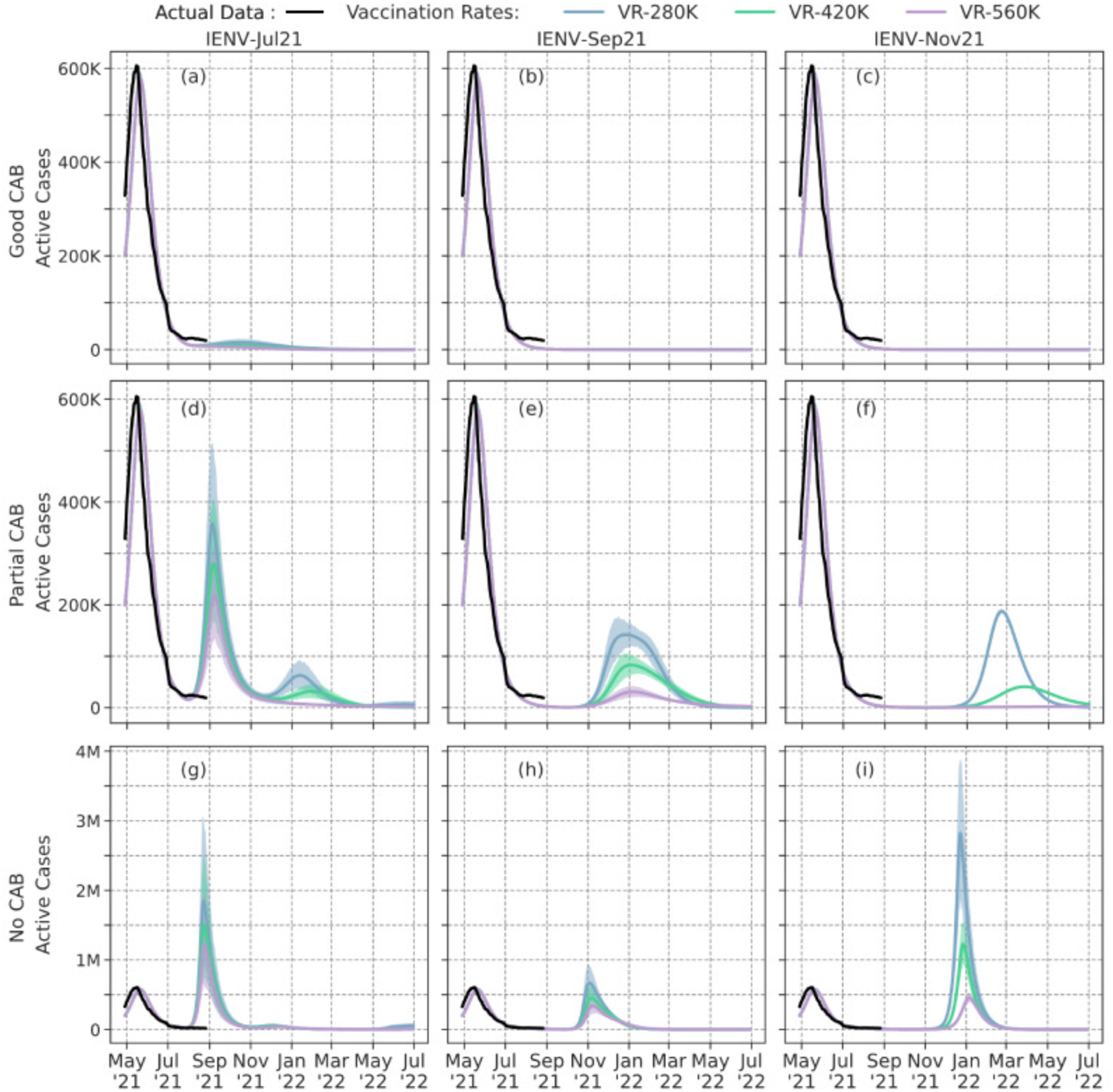
Time series of Mean of active cases with 95% Confidence Interval

These plots confirm the observations from Sect. 3.2, but now showing a time series of the active caseload distribution. As mentioned earlier, the scenarios with Partial CAB (middle row in Fig. 4) are likely to be followed due to the re-opening of economy including offices and educational institutions. The impact of the new wave will be more when it emerges early (Fig. 4 (d)). At the peak of the new wave, the active caseload will be very high with the present vaccination rate, around 350K, when a new variant emerges before September 2021. If we assume that there is no antibody waning at least for 180 days, then, a new wave is predicted only at the beginning of next year, even if a new variant emerges before September 2021. In this case, the impact will be minimal. The benefits of increasing the vaccination rate can clearly be seen in Fig. 4 (e). Though a new wave is predicted in a few scenarios, the active caseload is very minimal with the doubled vaccination rate. If a new variant does not emerge until the end of October 2021, then a new wave can be suppressed by doubling the vaccination rate (Fig. 4 (f)). In this case, the study shows that the active caseload will be as high as 200K with the present vaccination rate even if the new variant emerges after October 2021. This finding emphasizes the necessity of vaccinating the entire population by December 2021.

## 4 Discussion and Summary

We have undertaken a comprehensive study by quantifying several uncertain causal factors to provide modeling evidence on the emergence of a new COVID wave in Karnataka, India. A total of 972 ensemble members were formed by varying seven key causal factors in our Population Balance PDE model for infectious disease spread [21]. This first-principles based model incorporates the nonlinear dynamics between all the causal factors and the confirmed, active, recovered, and deceased caseload of Covid-19 in Karnataka, India. The major findings of this study are:

1. A new wave is most sensitive to mutation (emergence of immune-escape new variant) timing, compliance of COVID Appropriate Behavior, and vaccination rate.
2. Practically, the critical causal factor is compliance to CAB, which can be regulated with appropriate NPI such as mobility restriction, masking and physical distancing mandates, and crowd control measures.
3. Emergence of new variants beyond Sep’21 is not likely to induce new waves when the social distancing is good (lockdown-like restrictions).
4. Among scenarios with Wave-3, increasing the vaccination rate reduces the peak active caseload and consequently decreases hospitalization, ICU and Oxygen requirements.
5. Scenarios with a vaccination rate of 480k per day (2x the average vaccination rate as on July 4, 2021) should have a lower probability of Wave-3 than other scenarios
6. The daily confirmed cases of children (Age 0-11 and 12-17 years) at peak could be on an average seven times more than the corresponding daily confirmed cases at Wave-2 peak.

The above findings raise an important call for action to the state’s public health response. First, policies to promote CAB so as to ensure at least partial compliance from the population is required. Next, increasing the vaccination rate so as to fully vaccinate the entire population by mid-Dec is necessary to counter new variants and antibody waning, if any. Based on the predicted increase in daily confirmed case load of children in a potential wave 3 compared to data of wave 2, the policymakers can plan pediatric COVID care centers.

Though the estimated Wave-3 numbers are for the state of Karnataka, the dynamics of the ensemble forecast will be applicable to all other states of India. Therefore, the above recommended policy interventions can be imposed nation-wide.

One limitation of the ensemble forecast is that we have not considered the possibility of new variants infecting the vaccinated population, but we have considered a fixed efficacy of 70% for the vaccinated population that can partially offset the limitation. Further, the recovery rate is fitted to the data from July 2020 to May 2021; however, it could be more due to vaccination effect or less due to new variants.

## Data Availability

Data is available on request

## Acknowledgments

We thank members of the Karnataka Technical Advisory Committee, and Dr. Vinod Paul of Niti Aayog, Govt. of India for insightful questions and discussion. All reported COVID data is obtained from www.covid19india.org. We are thankful to the MHRD Grant No. STARS-1/388 (SPADE) for partial support. Thivin is supported by a Graduate Fellowship from the Ministry of Education, Govt. of India. Deepak acknowledges the Arcot Ramachandran Young Investigator award for partial support.

